# Lesion covariance networks reveal proposed origins and pathways of diffuse gliomas

**DOI:** 10.1101/2021.04.09.21255209

**Authors:** Ayan S. Mandal, Rafael Romero-Garcia, Jakob Seidlitz, Michael G. Hart, Aaron Alexander-Bloch, John Suckling

## Abstract

Diffuse gliomas have been hypothesized to originate from neural stem cells in the subventricular zone. Here, we evaluated this hypothesis by mapping independent sources of glioma localization and determining their relationships with neurogenic niches, genetic markers, and large-scale connectivity networks. Using lesion data from a total of 410 patients with glioma, we identified -- and replicated in an independent sample -- three lesion covariance networks (LCNs), which reflect clusters of frequent glioma co-localization. Each LCN overlapped with a distinct horn of the lateral ventricles. The first LCN, which overlapped with the anterior horn, was associated with low-grade, IDH-mutated/1p19q-codeleted tumors, as well as a neural transcriptomic signature and improved overall survival. Each LCN significantly corresponded with multiple brain networks, with LCN1 bearing an especially strong relationship with structural and functional connectivity, consistent with its neural transcriptomic profile. Finally, we identified subcortical, periventricular structures with functional connectivity patterns to the cortex that significantly matched each LCN. Cumulatively, our findings support a model wherein periventricular brain connectivity guides tumor development.

## Introduction

Diffuse gliomas are among the most lethal forms of cancer, yet the etiology and pathogenesis of this condition is not well understood. A significant barrier to optimal treatment for gliomas is a lack of clarity regarding the anatomical origins and migration patterns of the tumors. In contrast to early ideas that gliomas originate from mature glial cells in the same locations where they are observed, current theories imply that tumors originate from neurogenic niches in the subventricular zone (SVZ), from which they then migrate to populate distributed brain areas (Jiang and Uhrbom, 2012; Lee et al., 2018; Sanai et al., 2005). This idea is supported by genomic evidence from patients (Lee et al., 2018) as well as the observation of significantly elevated glioma frequency surrounding neurogenic niches (Mandal et al., 2020). In parallel, other research has indicated that glioma stem cells travel along previously healthy brain structures, including blood vessels and white matter tracts, suggesting that large-scale connectivity networks may help describe glioma migration (Cuddapah et al., 2014; Gillespie and Monje, 2018). However, the pathways by which gliomas could progress from subventricular origins to their final destinations remain speculative.

The natural progression of other neurological diseases, such as frontotemporal dementias, Alzheimer’s disease, and Parkinson’s disease, has been most reliably investigated using longitudinal brain imaging of large cohorts of patients (Brown et al., 2019; Vogel et al., 2020; Yau et al., 2018). This approach is difficult to apply to brain tumors, which are typically treated only weeks after initial diagnosis. An alternative method for probing brain development and degeneration from cross-sectional data is the use of structural covariance analysis (Alexander-Bloch et al., 2013a). Structural covariance networks identify correlations in brain size (measured by cortical thickness or volume) between brain regions across large cohorts of healthy or diseased individuals (Mechelli et al., 2005). These interregional relationships reflect a range of shared biological influences, including coordinated development, connectivity, and genetic similarity (Alexander-Bloch et al., 2013b; Romero-Garcia et al., 2018; Yee et al., 2018).

Analogously, interregional correlations in brain atrophy within defined neurological syndromes have been demonstrated to reflect patterns of coordinated degeneration and network spread of pathology (Seeley et al., 2009). Pairs of brain regions which are both consistently affected by a neuropathology could have this relationship for a number of informative reasons, such as a shared biological vulnerability, or a common pathway along which the disease spreads (Vanasse et al., 2021). The latter possibility is supported by studies of Parkinson’s and Alzheimer’s disease, which have unveiled networks of brain atrophy and tau accumulation that follow intrinsic functional connectivity networks (Ossenkoppele et al., 2019; Zeighami et al., 2015). The notion that patterns of collateral damage can reveal insights into the etiology of brain disease is also supported by the phenomenon of lesion covariance in stroke, which stems from the vascular origins of the injury (Mah et al., 2014; Zhao et al., 2020). An analysis of the networks of brain regions which tend to be co-affected by glioma tumors, therefore, may reveal insights into the possible ventricular origins of these deadly brain cancers, and point to pathways by which the tumors spread throughout the brain.

In this study, we applied independent component analysis (ICA) to tumor masks of patients with low- and high-grade glioma to identify networks of brain regions co-lesioned by gliomas (i.e. lesion covariance networks (LCNs)). Next, we examined associations between these networks and clinically relevant patient information, such as tumor grade, molecular genetics, transcriptomic signature, and overall survival. Finally, we related the LCNs to large-scale functional and structural connectivity networks to identify the potential pathways that underlie tumor development. We hypothesized that LCNs would coincide with the three horns of the lateral ventricles, and that the connectivity patterns of periventricular brain regions would correspond with the observed cortical locations of the tumors.

## Results

### Lesion covariance networks implicate horns of the lateral ventricles

We applied ICA to spatially-aligned masks of tumor volume derived from a validated imaging processing pipeline applied to presurgical brain MRIs of 242 high-grade and low-grade glioma patients (Bakas et al., 2017). ICA identified three independent components with scores across patients and voxels (Figure 1A). Given the similarity of the methodological approach to functional connectivity and structural covariance analyses, we decided to refer to the resulting independent components as lesion covariance networks.

**Figure 1.**
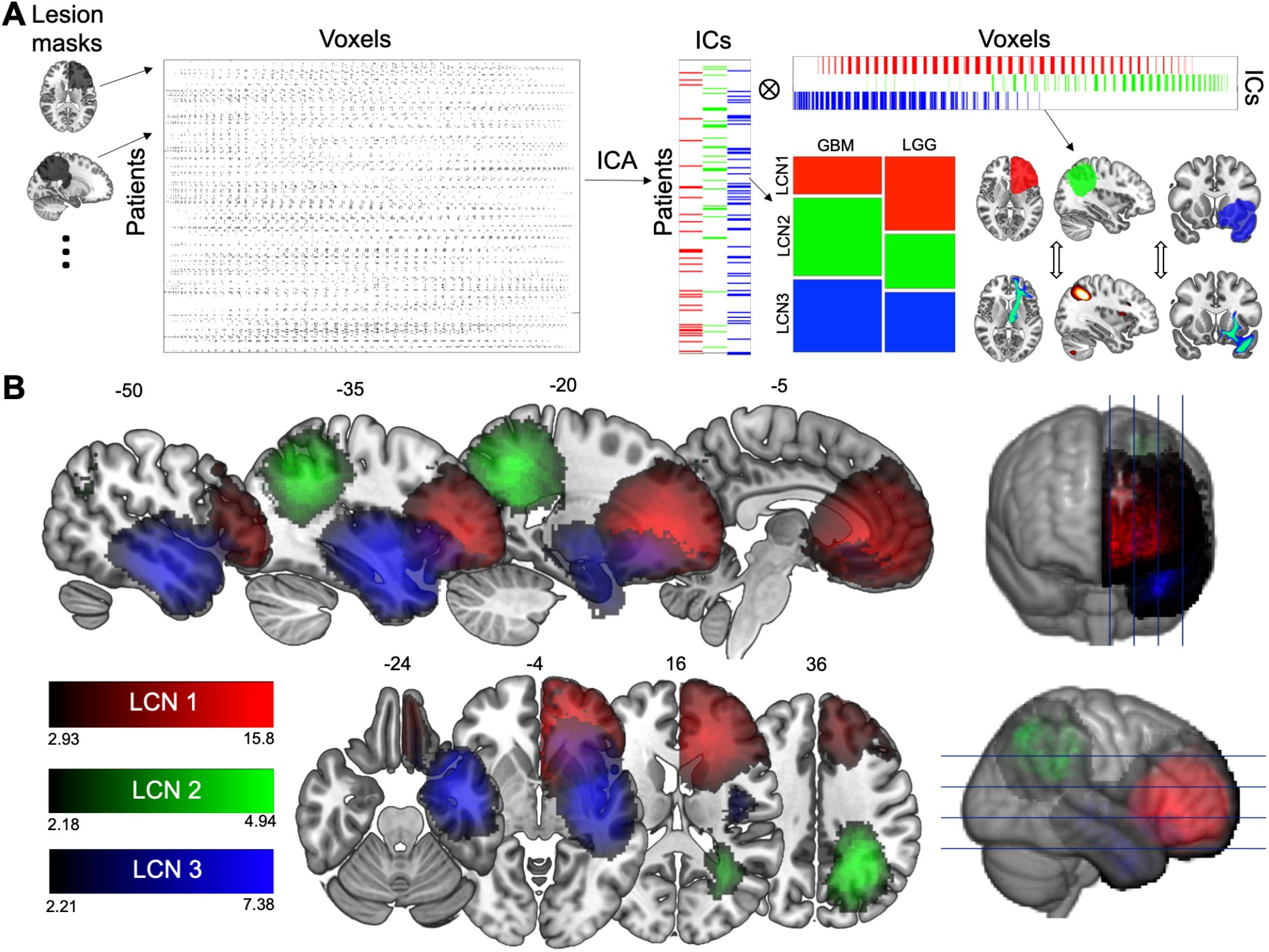
Lesion covariance networks of glioma localization revealed by independent component analysis. (A) Overview of the study. Lesion masks from 242 glioma patients were mapped to one hemisphere then concatenated to form a voxel-wise matrix. This matrix was decomposed via ICA into i) IC scores, which were related to clinical variables, and ii) spatial maps (i.e. LCNs), which were cross-correlated with structural and functional connectivity networks. (B) LCNs are displayed, thresholded to include positive voxels with over 50% likelihood of association with the independent component. Abbreviations: LCN = lesion covariance network; ICA = independent component analysis; IC = independent component; GBM = glioblastoma; LGG = low-grade glioma.

ICA revealed three LCNs which extended into the frontal, parietal, and temporal lobes respectively (Figure 1B). Notably, each LCN overlapped with a distinct horn of the lateral ventricles, with LCN1 covering the anterior horn, LCN2 covering the posterior horn, and LCN3 covering the inferior horn. The same LCN locations were replicated in an independent set of 168 glioma patients (Supplementary Figure 1).

### Clinical outcomes associated with LCNs

To determine how the LCNs related to important clinical variables such as cellular pathology and molecular genetics derived from the same patient sample, we first assigned each patient to one of three groups based on the LCN with which their tumor was most associated. Chi-square tests indicated significant associations between LCN group and tumor grade (χ^2^(2) = 11.1; *p* = 0.0038), as well as between LCN group and IDH/1p19q-status (χ^2^(2) =6.7; *p* = 0.03) (Figure 2A). Post-hoc tests with Bonferroni correction indicated that LCN1 was significantly overrepresented in low-grade gliomas (LGG; Pearson residuals = 3.29; *p* = 0.0059) and IDH-mutated/1p19q-codeleted tumors (residuals = 5.65; *p* < 1e-6), but underrepresented in glioblastoma (GBM; residuals = −3.29; *p* = 0.0059) and IDH-wildtype tumors (residuals = −4.05; *p* = 0.00046). LCN2 was positively associated with IDH-wildtype tumors (residuals = 2.84; *p* = 0.04), whereas LCN3 was negatively associated with IDH-mutated/1p19q codeleted tumors (residuals = −3.39; *p* = 0.006). These results implicate LCN1 as a potential radiological signature of IDH-mutated/1p19q-codeleted status, which is pathognomonic of oligodendroglioma.

**Figure 2.**
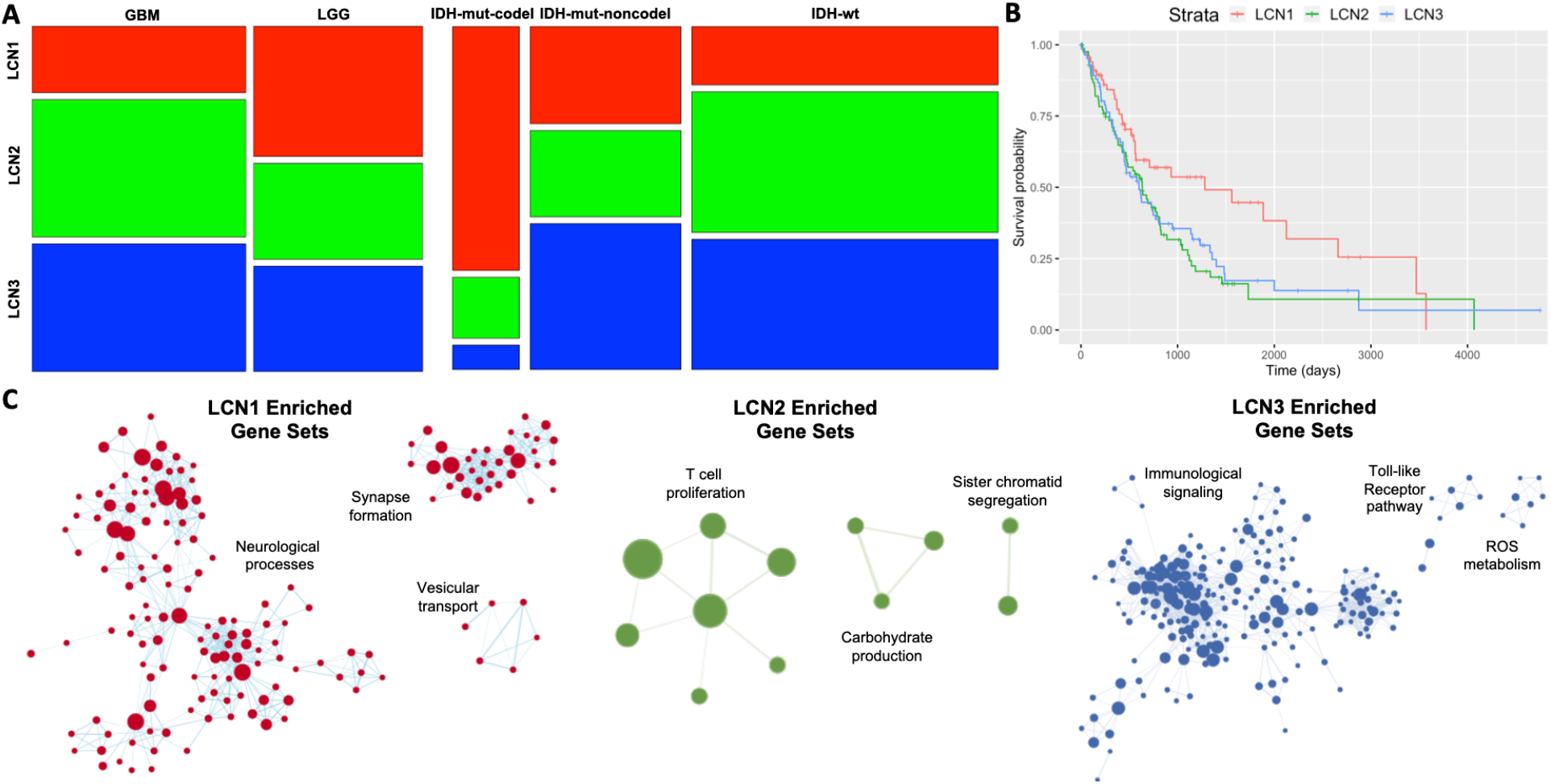
Clinical and genomic correlates of lesion covariance networks. (A) Mosaic plots represent the proportion of gliomas within each LCN associated with clinical variables, including tumor grade and molecular subtype. (B) Kaplan-Meier curves show overall survival outcomes stratified by LCN group. (C) Gene ontology networks associated with differentially-expressed genes for each LCN. Enriched gene sets are plotted as nodes, with gene set size proportional to node size, and the similarities between gene sets are represented as edges. Network components with the three highest numbers of nodes are displayed.

Next, we related the LCNs to overall survival, first by visualizing Kaplan-Meier curves stratified by LCN group (Figure 2B). Patients in the LCN1 group had notably prolonged survival compared to patients in the other groups. This association was confirmed statistically and shown to be independent of potential confounding demographic variables through a Cox Proportional Hazards regression model (Table 1). Interestingly, the effect of LCN1 was no longer significant after clinical variables (i.e. tumor grade and IDH/1p19q-status) were included in the model, suggesting that the association between LCN group and tumor molecular genetics drove the differences in survival outcome.

**Table 1.**
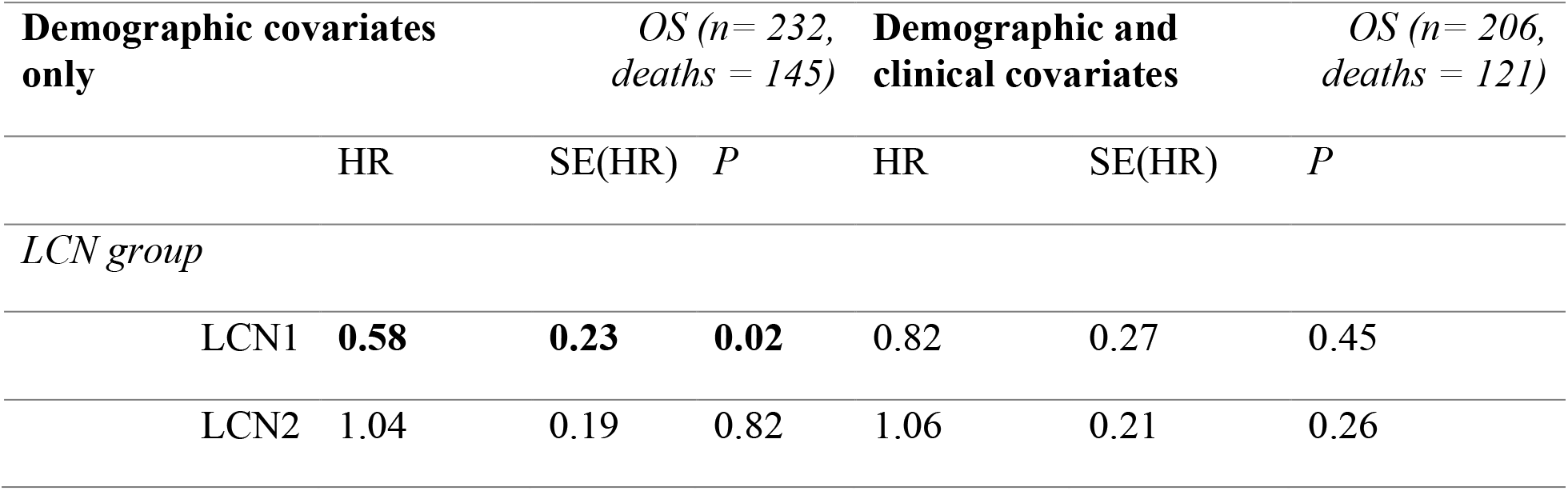
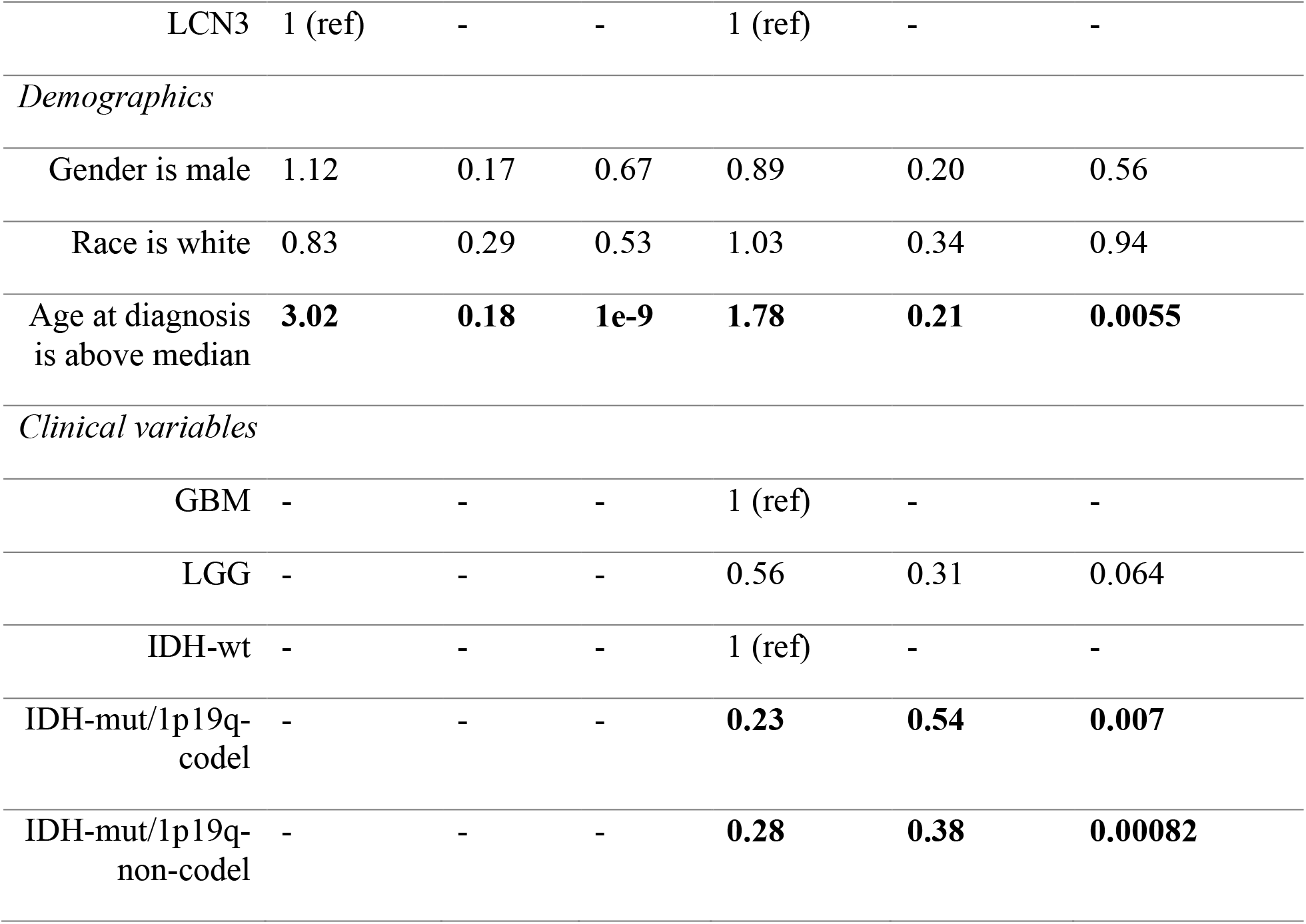
Cox Proportional Hazards models relating LCN group and demographic/clinical covariates with overall survival. Abbreviations: OS=overall survival; HR=hazards ratio; SE=standard error.

| Demographic covariates only |  | OS (*n* = 232, deaths = 145) |  |  | Demographic and clinical covariates |  | OS (*n* = 206, deaths = 121) |
| --- | --- | --- | --- | --- | --- | --- | --- |
|  | HR | SE(HR) | *P* |  | HR | SE(HR) | *P* |
| *LCN group* |  |  |  |  |  |  |  |
| LCN1 | **0.58** | **0.23** | **0.02** |  | 0.82 | 0.27 | 0.45 |
| LCN2 | 1.04 | 0.19 | 0.82 |  | 1.06 | 0.21 | 0.26 |

|  |  |  |  |  |  |  |
| --- | --- | --- | --- | --- | --- | --- |
| LCN3 | 1 (ref) | - | - | 1 (ref) | - | - |
| *Demographics* |  |  |  |  |  |  |
| Gender is male | 1.12 | 0.17 | 0.67 | 0.89 | 0.20 | 0.56 |
| Race is white | 0.83 | 0.29 | 0.53 | 1.03 | 0.34 | 0.94 |
| Age at diagnosis is above median | **3.02** | **0.18** | **1e-9** | **1.78** | **0.21** | **0.0055** |
| *Clinical variables* |  |  |  |  |  |  |
| GBM | - | - | - | 1 (ref) | - | - |
| LGG | - | - | - | 0.56 | 0.31 | 0.064 |
| IDH-wt | - | - | - | 1 (ref) | - | - |
| IDH-mut/1p19q-codel | - | - | - | **0.23** | **0.54** | **0.007** |
| IDH-mut/1p19q-non-codel | - | - | - | **0.28** | **0.38** | **0.00082** |

### LCN groups diverge in their expression of neural versus inflammatory genes

In a subset of patients for whom bulk RNA-sequencing data were available, we performed differential expression analyses to identify genes upregulated among the primary tumors of each LCN group relative to the other groups. Gene set enrichment analyses were performed on the resulting ranked gene lists, and the enriched ontologies were visualized as a network (Figure 2C). LCN1 was positively enriched with a large number of gene sets associated with neurological processes, such as synaptic signaling and cognition, as well as ontologies involving synapse formation and vesicular transport. The largest network components, comprised of gene sets enriched in LCN2 and LCN3, related respectively to T cell proliferation and immunological signaling. Given previous reports of synaptic enrichment among low-grade tumors and oligodendrogliomas (Ceccarelli et al., 2016; Venkatesh et al., 2019), we performed a follow-up gene set enrichment analysis for LCN1 where tumor grade and IDH/1p19q status were included as covariates, and found that LCN1 remained enriched for synaptic signaling (Supplementary Figure 2).

### LCNs relate to large-scale connectivity networks

Next, we assessed anatomical correspondence between LCNs and large-scale connectivity networks, by correlating the three LCNs with 21 functional connectivity networks and 11 white matter pathways derived from a large, healthy neuroimaging dataset (Miller et al., 2016). LCN1 significantly corresponded with four functional connectivity networks each with strong frontal components, including the cingulo-opercular, anterior salience, dorsal attention, and frontoparietal networks. LCN1 also corresponded to two major white matter pathways: the anterior thalamic radiation and the uncinate fasciculus. LCN2 corresponded with two white matter pathways and two functional networks, including the posterior default mode network, whereas LCN3 corresponded with the auditory network. Effect sizes and p-values for all connectivity networks that were statistically significant after correction for multiple comparisons are shown in Supplementary Table 3. The strongest correspondences between LCNs and large-scale connectivity networks are displayed in Figure 3A, with other significant associations are shown in Supplementary Figure 3.

**Figure 3.**
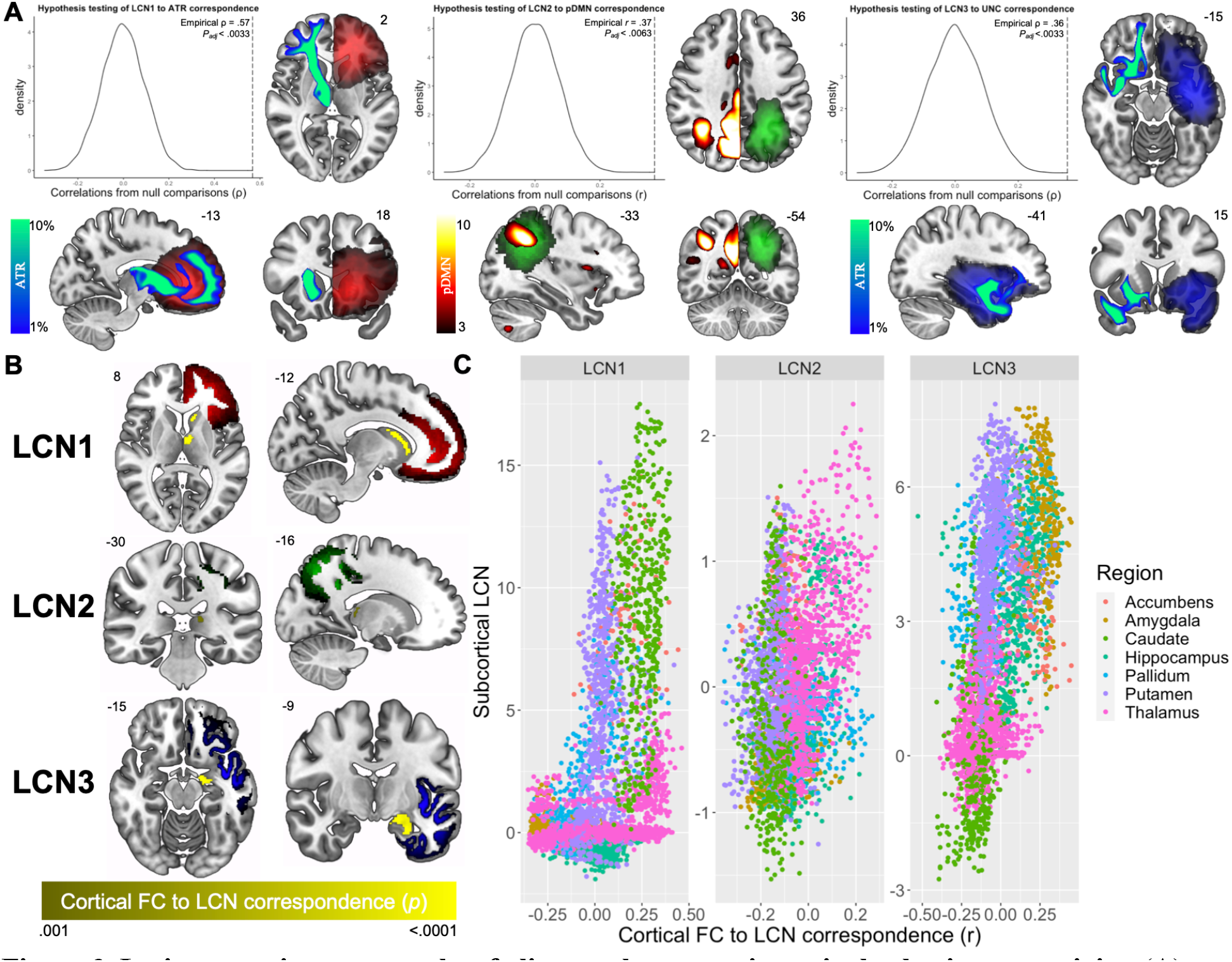
Lesion covariance networks of glioma relate to periventricular brain connectivity. (A) Structural and functional connectivity networks with the strongest correlation with each LCN. Significance of correspondence was assessed by comparison with spatial-autocorrelation-preserving surrogate LCN maps (Burt et al., 2020). LCNs are colored with the same scale as in Figure 1. Structural connectivity networks (where streamline density is represented by a winter color scale) and functional connectivity networks (where connectivity strength is represented by a hot color scale) are displayed on the opposite hemisphere of the LCN for visualization in axial and coronal slices. See Supplementary Figure 3 for other significantly associated connectivity networks. (B) Subcortical voxels are colored based on the significance of the association between their seed-based functional connectivity map and the cortical values of each LCN map (voxel-wise *P* < .001; cluster-level *P* < .05). The LCNs are also shown with the same color scale as in Figure 1. (C) Scatterplots illustrate subcortical structures with both high functional correspondence and involvement with each LCN, found in the upper right quadrant of each plot. Abbreviations: ATR = anterior thalamic radiation, pDMN = posterior default mode network, UNC = uncinate fasciculus.

#### Cortical LCN locations match functional connectivity with periventricular brain areas

Finally, to identify subcortical structures that may drive the correspondence between connectivity and cortical lesion location, we performed seed-based functional connectivity analyses with all subcortical gray matter voxels and correlated the resulting maps with each LCN. We identified subcortical, periventricular clusters of voxels with cortical functional connectivity patterns that significantly matched each LCN (voxel-wise *P* < 0.001, cluster-level *P* < 0.05; Figure 3B). To determine the particular structures driving the observed relationships, we generated a scatterplot to highlight subcortical structures with both high functional correspondence and involvement with the three LCNs (Figure 3C), implicating the caudate, thalamus, and amygdala respectively.

## Discussion

In this study, we demonstrated replicable patterns of glioma localization with clinical relevance and spatial correspondence with large-scale functional and structural connectivity networks. Our findings provide evidence for the subventricular origins of glioma, delineate an imaging signature linked to tumor genetics, and contribute to a growing literature on the bidirectional relationship between gliomas and their neural microenvironment.

### Subventricular origins of glioma

Contact with the lateral ventricles is a known prognostic factor for gliomas, predicting poorer overall survival (Chaichana et al., 2008; Jafri et al., 2013; Mistry et al., 2017a, 2017b) and tumor recurrence (Adeberg et al., 2014; Chen et al., 2015). These observations have motivated the popular notion that neurogenic niches of the SVZ act as a tumor reservoir, contributing to the therapeutic resistance of diffuse gliomas (Altmann et al., 2019). However, there remains a point of debate as to whether gliomas spread to the SVZ or if the tumor originates in this area. Our findings inform this debate by establishing: (1) that gliomas cluster around the horns of the lateral ventricles; and (2) that connectivity with periventricular regions corresponds significantly with cortical tumor locations. Our results are most consistent with a model where gliomas originate in any of the three horns of the lateral ventricles, then migrate along neural pathways to arrive at their cortical destinations. The results also suggest that for cases of glioma that appear from radiological imaging to spare the SVZ, the SVZ may nevertheless harbor oncogenic stem cells. This conclusion is consistent with a recent study that found cancer-driving mutations in radiologically tumor-free SVZ tissue in glioblastoma patients (Lee et al., 2018).

While our statistical decomposition of glioma distribution captures the lesion locations common to the majority of these tumors, it is worth noting that there are gliomas which do not fit the patterns of any of the lesion covariance networks described in this study. For example, none of the LCNs substantially covered the supplementary or primary motor areas, where gliomas are sometimes observed. These tumors are often positioned directly above the central body of the lateral ventricles, so one could speculate that the gliomas originate there. Future work could decompose glioma distribution with a higher dimensionality in order to more comprehensively characterize glioma location patterns and identify their sources.

### A radiological signature for oligodendrogliomas

The accurate prediction of molecular genetic subtype from tumor imaging is a crucial goal of the burgeoning field of radiogenomics (Ellingson, 2015; Fathi Kazerooni et al., 2019) and a potentially transformative clinical tool to aid early and precise glioma diagnosis. Research in this area has illustrated important associations between tumor location and molecular genetic signatures, the most robust of these being a propensity for IDH-mutated gliomas to localize to the rostral end of lateral ventricles (Tejada Neyra et al., 2018; Wang et al., 2015). However, the previous studies in this area used voxel-based lesion symptom mapping, an approach that necessitates stringent multiple comparisons correction (Mirman et al., 2018), thereby reducing their power to detect localization differences between subtypes of IDH-mutated tumors. We limited the number of statistical comparisons involved in our study by first reducing the dimensionality of the lesion data. As a result, we were able to reveal a specific association between IDH-mutated/1p19q-codeleted status (pathognomonic of an oligodendroglioma tumor) and lesion location in frontal cortex. This result is consistent with reports from other, more qualitative neuroimaging studies investigating oligodendroglioma localization (Ellingson, 2015; Zlatescu et al., 2001).

Our findings also suggest that tumors with different molecular genetic signatures preferentially arise from different portions of the ventricular lining. A possible explanation for this result is that some gliomas may need a specific metabolic niche in order to thrive and develop into a symptomatic brain tumor. Consistent with this idea, some studies have proposed that high glutamate flux, and the restricted expression of hominoid-specific glutamate dehydrogenase enzymes in the prefrontal cortex, support the survival of IDH-mutant glioma cells in this region (Chen et al., 2014; Waitkus et al., 2018). Symbiotic relationships between cancer cells and the tumor microenvironment, often framed within the “seed and soil hypothesis”, have helped explain the metastatic patterns of other cancers and likely also bear relevance for glioma development.

### Network spread of glioma tumors

Migration of glioma tumors along pre-existing brain structures, including blood vessels and white matter tracts, has been acknowledged since the 1930s (Scherer, 1938, 1940). Over the last decade, it has been demonstrated that migration along these structures is not simply a stochastic process by which tumors follow paths of least resistance (Cuddapah et al., 2014). Rather, glioma cell migration is coordinated, in part, by signaling molecules secreted during neuronal firing, in a process of activity-dependent glial cell proliferation that is also a key mechanism in healthy brain development (Gillespie and Monje, 2018; Venkatesh and Monje, 2017; Venkatesh et al., 2019). Our findings contribute to this literature by illustrating, in human patients, that glioma localization follows intrinsic functional and structural connectivity networks. This result is consistent with prior work from our group demonstrating that gliomas localize to functional brain hubs (Mandal et al., 2020).

Early studies of glioma development noted differences between tumor subtypes in their tendency to grow along pre-existing brain structures (Scherer, 1938). Therefore, some glioma subtypes may be expected to follow brain connectivity networks more closely than others. Our study found support for this idea in that LCN1 related to eight connectivity networks, while the other two LCNs related to just two and four connectivity networks, respectively. Moreover, in addition to possessing the strongest correspondence to brain connectivity, LCN1 was also enriched with genes involved in neuronal processes such as synaptic signaling and synapse formation. A recent study illustrated that glioma cells enriched with these types of genes integrate into neural circuits involved in lexical processing, and that this neural integration supports tumor proliferation (Krishna et al., 2021). The genomic signature of a glioma may thus be an important predictor of the tumor’s eventual migration patterns.

We interpret the correspondence between functional connectivity and glioma localization to reflect tumor migration along neuronal networks that support glioma cell proliferation. However, neuronal networks are known to relate intimately with the brain’s vasculature, which has been noted to be a critical spreading substrate for gliomas (Montana and Sontheimer, 2011). Neuronal and vascular networks converge on similar anatomy in adults (Bright et al., 2020), potentially reflecting the synergistic growth of neuronal and vascular processes during development (Quaegebeur et al., 2011; Wälchli et al., 2015). Therefore, one possible interpretation of our results is that the functional networks serve as a proxy for regions with common vascular inputs, and that gliomas invade these territories along the vasculature. This possibility is not mutually exclusive with our primary interpretation, given that neuronal activity could still be driving the migration along blood vessels. The exact physical substrate of activity-dependent glioma cell migration should be investigated further.

## Conclusion

A better understanding of the origins and migration patterns of gliomas could inform surgical and radiation treatments intended to comprehensively obliterate tumor cells. We demonstrated that gliomas cluster around distinct horns of the lateral ventricles, and that these tumor distribution patterns relate to diagnostic genomic signatures and large-scale connectivity networks. Our study connects two separate literatures on the subventricular origins of glioma and symbiotic glioma-neuron relationships to propose a model wherein periventricular brain connectivity guides glioma development.

## Methods

### Construction of lesion covariance networks

Neuroimaging data of patients with low- and high-grade glioma were accessed from The Cancer Imaging Archive (TCIA; www.cancerimagingarchive.net) (Bakas et al., 2017; Clark et al., 2013). Pre-operative multimodal (i.e. T1w, T1w-Gd, T2w, T2w-FLAIR) scans were obtained from 135 patients with high-grade gliomas and 108 patients with low-grade gliomas at thirteen institutions with different imaging sequences and protocols. These scans were skull-stripped, co-registered, and resampled to 1mm^3^ voxel resolution before being entered into GLISTRboost (Bakas et al., 2016), a top-ranked tumor segmentation algorithm, which classified voxels into four classes: contrast-enhancing tumor, necrotic non-enhancing core, peritumoral edema, and normal brain tissue. Labels were then manually corrected by board-certified neuroradiologists.To limit our study to supratentorial lesions, we removed one subject with a posterior fossa tumor. All patients were diagnosed with a diffuse glioma of grade II or higher. Demographic information of the patient sample is included in Supplementary Table 1.

T1-weighted images from each patient were nonlinearly warped to the Montreal Neurological Institute (MNI) 152 template using Advanced Normalization Tools (ANTS) software, with cost-function masking of abnormal brain tissue. Registered masks corresponding to contrast-enhancing tumor and the non-enhancing core were taken to represent, and henceforth will be referred to as, the tumor mask. To reduce the dimensionality of the data, each tumor mask in the right hemisphere was flipped to the left hemisphere, such that each mask was aligned to the same cerebral hemisphere.

Tumor masks from the 242 subjects were combined into one 4D data structure and then entered into Melodic Independent Component Analysis (ICA) in FSL (Jenkinson et al., 2012; Smith et al., 2004). ICA is a source separation algorithm that decomposes a dataset into a fixed number of statistically independent components (ICs). Given our hypothesis that tumors would stem from the anterior, posterior, and inferior horns of the lateral ventricles, we selected a dimensionality of three. The output of ICA included three brain maps of Z statistics indicating the likelihood of each voxel belonging to the corresponding IC, as well as three vectors indicating a score for each patient representing their tumor’s spatial association with each IC. Probabilistic maps for each lesion covariance network were generated using a mixture modeling approach (Beckmann and Smith, 2004), and were used to threshold each IC map at 0.5, excluding voxels with a higher likelihood of belonging to a background noise class than to the IC.

### Replication in an independent sample

To determine whether the lesion covariance networks identified in the TCIA dataset could be replicated, we performed ICA on images from the Brain Tumor Segmentation (BraTS) 2019 dataset (Bakas et al., 2017, 2018; Menze et al., 2015). This dataset includes manually segmented images from 335 low- and high-grade glioma patients, pre-processed in the same way as the TCIA dataset described above. We removed patients who overlapped between the two datasets, resulting in 168 subjects. The resulting spatial maps from ICA with three dimensions were cross-correlated with the LCNs from the TCIA dataset, and are displayed in Supplementary Figure 2.

### Relating LCNs to clinical variables

To determine how the LCNs related to clinically relevant information from the same cohort – including tumor grade, molecular genetics, and overall survival – we first assigned each patient to one of three groups based on the LCN for which their tumor had the highest IC score (i.e. the LCN with which their tumor was most associated). Then, we used chi-square tests to assess the association between these location-based groups and clinical variables such as tumor grade (GBM vs LGG) and molecular genetic subtype (IDH-wildtype vs IDH-mutant/1p19q codeletion vs IDH-mutant/1p19q non-codeletion). To compare overall survival outcomes between individuals in each LCN group, we plotted a Kaplan-Meier curve. Finally, we performed Cox-Proportional Hazards regressions to quantitatively assess the relationship between LCN group and overall survival. Two models were considered: the first model included LCN group (with LCN3 as the reference level) and demographic covariates (gender and age, binned by the median); the second model included LCN group, demographic covariates, and clinical variables (tumor grade and molecular genetic subtype). Clinical and demographic data were accessed from The Cancer Genome Atlas (TCGA) (McLendon et al., 2008; Silva et al., 2016; 2015). For each analysis, patients with missing data were excluded, resulting in different sample sizes for different tests.

### Bulk transcriptomic analyses

To determine if tumors corresponding to different LCNs possessed distinct transcriptomic signatures, we performed bulk RNA sequencing analyses to relate LCN groups to differential gene expression. Following a previously reported workflow (Silva et al., 2016), we downloaded 516 LGG and 155 GBM primary solid tumor samples, 106 and 29 of which could be matched to MRI scans we had for the TCIA-LGG and TCIA-GBM datasets respectively. To remove potential outliers, we performed an Array-Array intensity correlation, which resulted in a square matrix denoting the Pearson correlation across genes between each TCGA sample (GBM and LGG). No samples were removed after applying a previously established correlation threshold (*r*>0.6). Next, we normalized our RNA-seq data using the EDAseq package, implementing: (1) within-lane normalization to adjust for the effects of GC-content on read counts; (2) loess robust local regression, global scaling, and full quantile normalization (Risso et al., 2011); and (3) between-lane normalization to adjust for differences between lanes, such as sequencing depth. Finally, we filtered out mRNA transcripts with a quantile mean threshold of 0.25 across all samples, reducing the number of genes considered from 19866 to 14899.

Using the edgeR package (Robinson et al., 2009), we ran three differential expression analyses comparing between patients included and not included in each LCN group (e.g. LCN1 vs LCN2 and LCN3, etc.). Negative binomial generalized linear models were fit with tagwise dispersion estimated. For each differential expression analysis, genes were ranked by their log2 fold-changes, then entered into Gene Set Enrichment Analysis (GSEA) to find enriched gene sets associated with each LCN (Subramanian et al., 2005). Using Cytoscape and Enrichment Map (Merico et al., 2010; Shannon et al., 2003), GSEA results were displayed as an annotation module network, where enriched gene sets are plotted as nodes and the similarity between gene sets is represented as edges. Because gene sets downregulated for one LCN tended to be upregulated in another, we only plotted positively enriched gene ontologies.

### Connectivity analyses

We hypothesized that the LCNs would relate to large-scale functional and connectivity networks involved in guiding the development of the tumors. We accessed 21 functional connectivity and 12 structural connectivity networks derived from UK BioBank neuroimaging data of over 4000 neurologically healthy individuals (Miller et al., 2016). Functional connectivity networks were identified from a 25-dimensional ICA decomposition of resting-state fMRI data. Four non-neuronally driven components were excluded. Structural networks were identified using XTract, an automated tractography protocol to identify major white matter pathways with standardized seed, exclusion, and termination masks (Warrington et al., 2020). We considered four association fibers (inferior fronto-occipital fasciculus, uncinate fasciculus, inferior and superior longitudinal fasciculus), five projection fibers (acoustic radiation, corticospinal tract, anterior, posterior, and superior thalamic radiations), and two limbic fibers (cingulum, main part and hippocampal part). Each of these tracts have left and right counterparts; therefore, streamline density maps from the left and right tracts were aligned to the same hemisphere and averaged. Functional connectivity networks, which also present bilaterally, were similarly mapped to one hemisphere and averaged.

The correspondence between LCNs and structural connectivity networks was quantified by calculating a voxel-wise Spearman’s rank correlation between maps, whereas LCN and functional connectivity correspondence was assessed using Pearson’s correlations. The statistical significance of brain map correspondence was determined by comparing the empirical correlation coefficient with coefficients derived from correlations with 10,000 spatial-autocorrelation-preserving surrogate LCN maps generated by BrainSMASH (Burt et al., 2020). This approach addresses the important confound of spatial autocorrelation to allow for an accurate P-value estimation. For the comparisons between LCNs and functional connectivity, we correlated values corresponding to the cortical and subcortical gray matter voxels of each brain map. Comparisons between LCNs and structural connectivity involved voxels with greater than 1% of the total number of streamlines identified by the XTract protocol.

Finally, we systematically performed seed-based functional connectivity (SBFC) analyses with each subcortical gray matter voxel and correlated the resulting maps with each LCN to identify structures that drive the correspondence between connectivity and lesion covariance. For each voxel in the Harvard-Oxford Subcortical Atlas, we calculated functional connectivity between the subcortical voxel and each cortical gray matter voxel, using the principal components of the UK BioBank Dense Functional Connectome (Smith et al., 2014; https://www.fmrib.ox.ac.uk/ukbiobank/). The resulting cortical SBFC maps were then normalized using the Fisher Z-transformation, and smoothed at 5 mm full-width half maximum. For each LCN, *P*-values were assigned to each subcortical voxel based on the significance of the relationship between its SBFC map and the LCN map using BrainSMASH. The three resulting subcortical *P*-maps were then thresholded to control for multiple comparisons (voxel-wise *P* value < 0.001; cluster-level family-wise error corrected *P* value < 0.05).

**Supplementary Table 1.** Demographic, clinical, and imaging variables for 242 patients with glioma from The Cancer Imaging Archive. When applicable, the standard deviation is in parentheses. Abbreviations: M=male; F=female; NA=not applicable; GBM=glioblastoma; LGG=low-grade glioma; IDH=isocitrate dehydrogenase; LCN=lesion covariance network.

|  |  |
| --- | --- |
| ***Demographic variables*** |  |
| Age (years) | 52.9 (15.2) |
| Gender (M/F/NA) | 133/107/2 |
| Race (white/non-white/NA) | 210/23/9 |
| ***Clinical variables*** |  |
| Grade (GBM/LGG) | 135/107 |
| Molecular subtype (IDH-wt/IDH-mut-1p19q-codel/IDH-mut-1p19q-noncodel/NA) | 124/27/61/30 |
| **LCN groups** |  |
| LCN groups (LCN1/LCN2/LCN3) | 69/87/86 |

**Supplementary Table 2.**
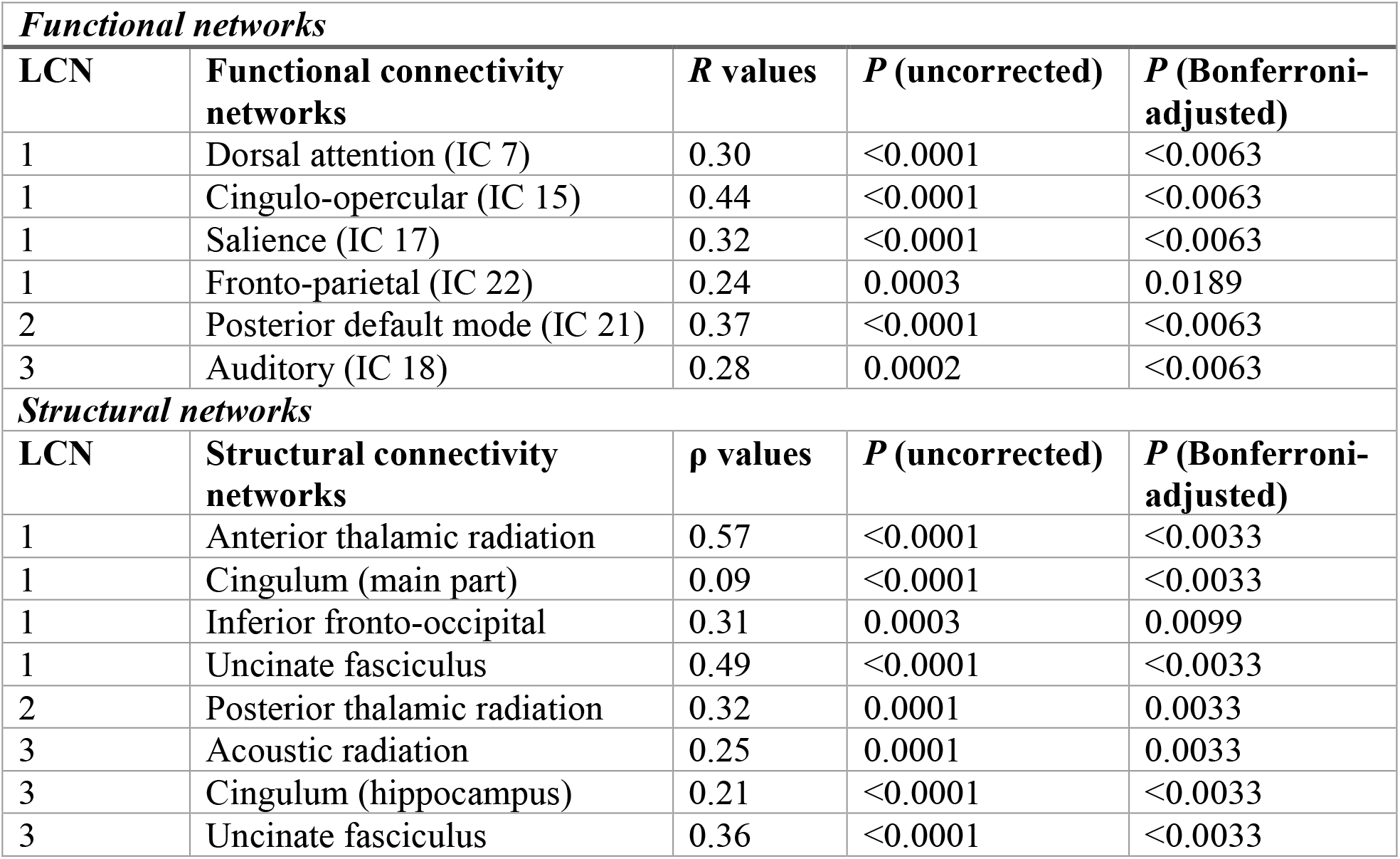
Functional and structural connectivity networks with significant correspondence to lesion covariance networks.

**Supplementary Figure 1.**
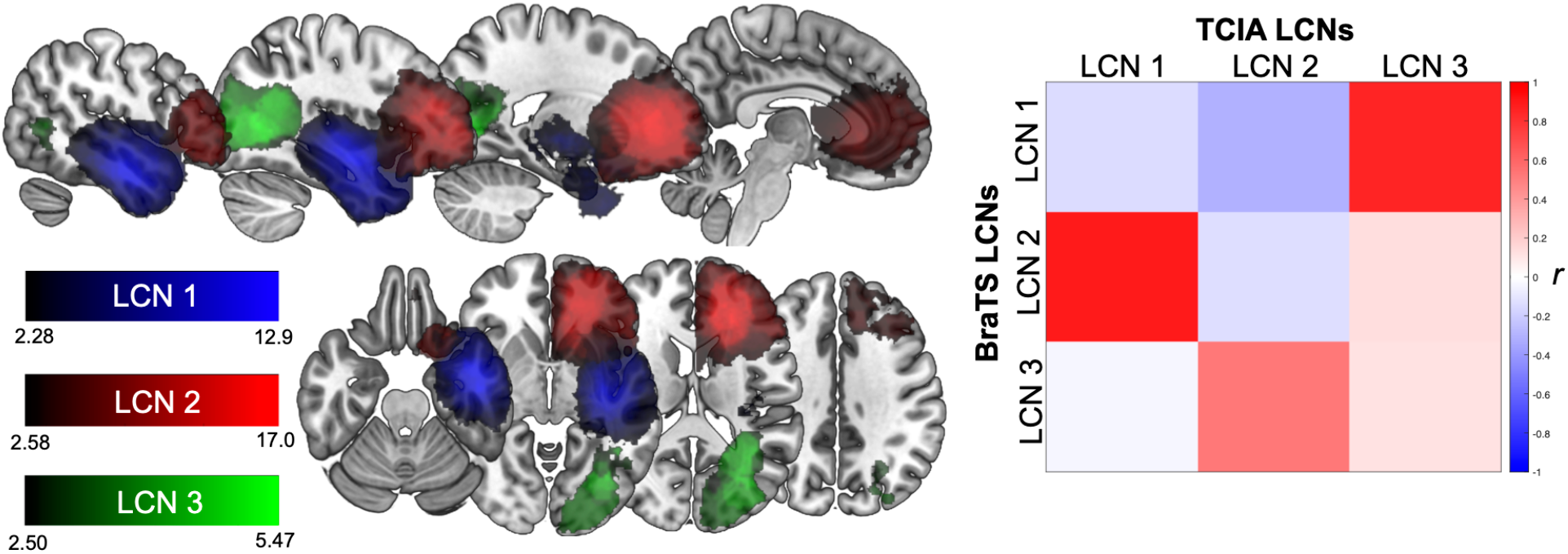
Replication of lesion covariance networks in an independent cohort. (A) LCNs derived from a cohort of 168 high- and low-grade glioma patients displayed on the same slices shown in Figure 1. (B) Correlation matrix illustrating the correspondence between LCNs in the TCIA and BraTS cohorts. Abbreviations: TCIA = The Cancer Imaging Archive; BraTS = Brain Tumor Segmentation Challenge.

**Supplementary Figure 2.**
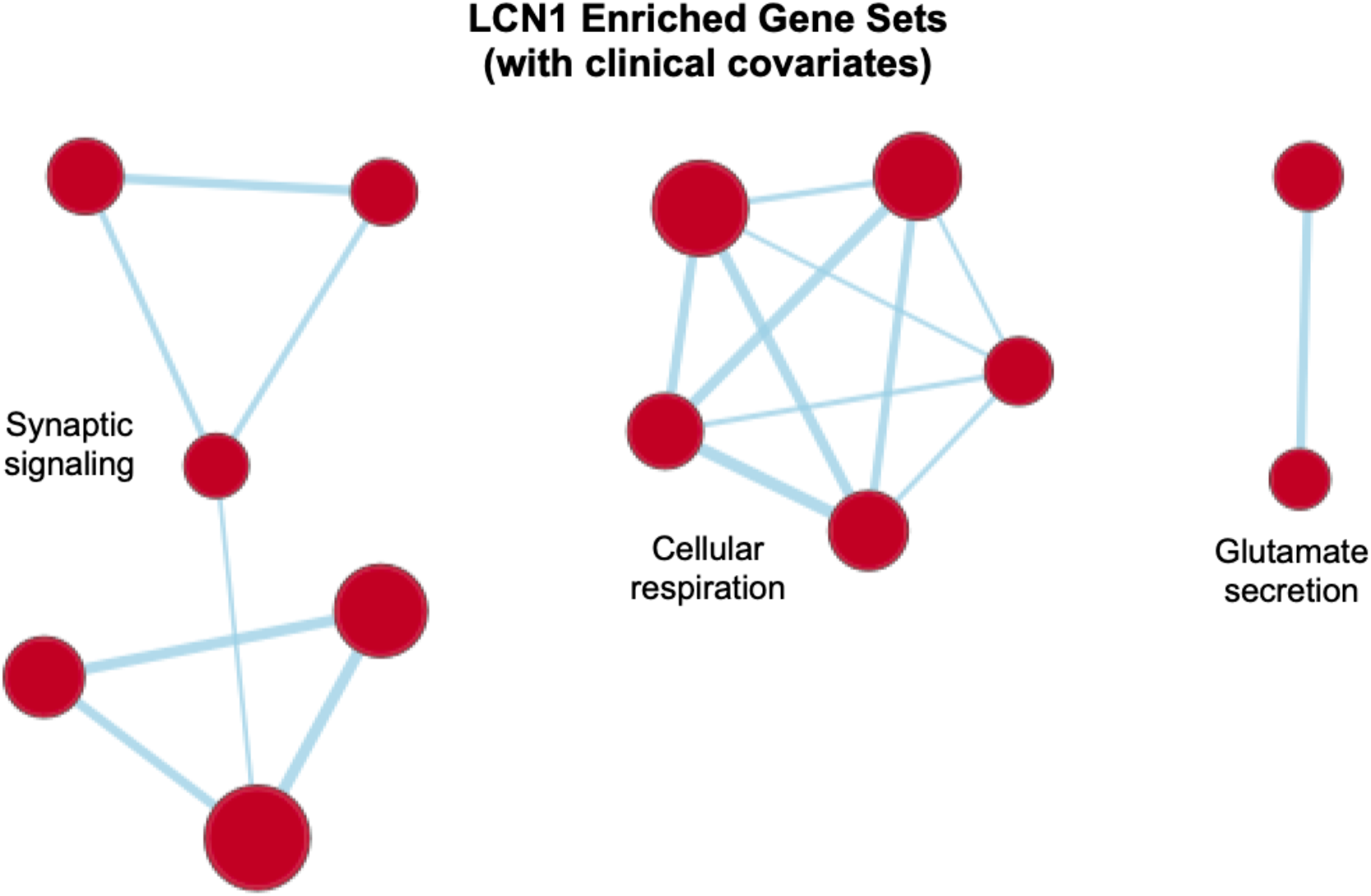
Gene ontology networks associated with LCN1 after controlling for tumor grade and molecular subtype.

**Supplementary Figure 3.**
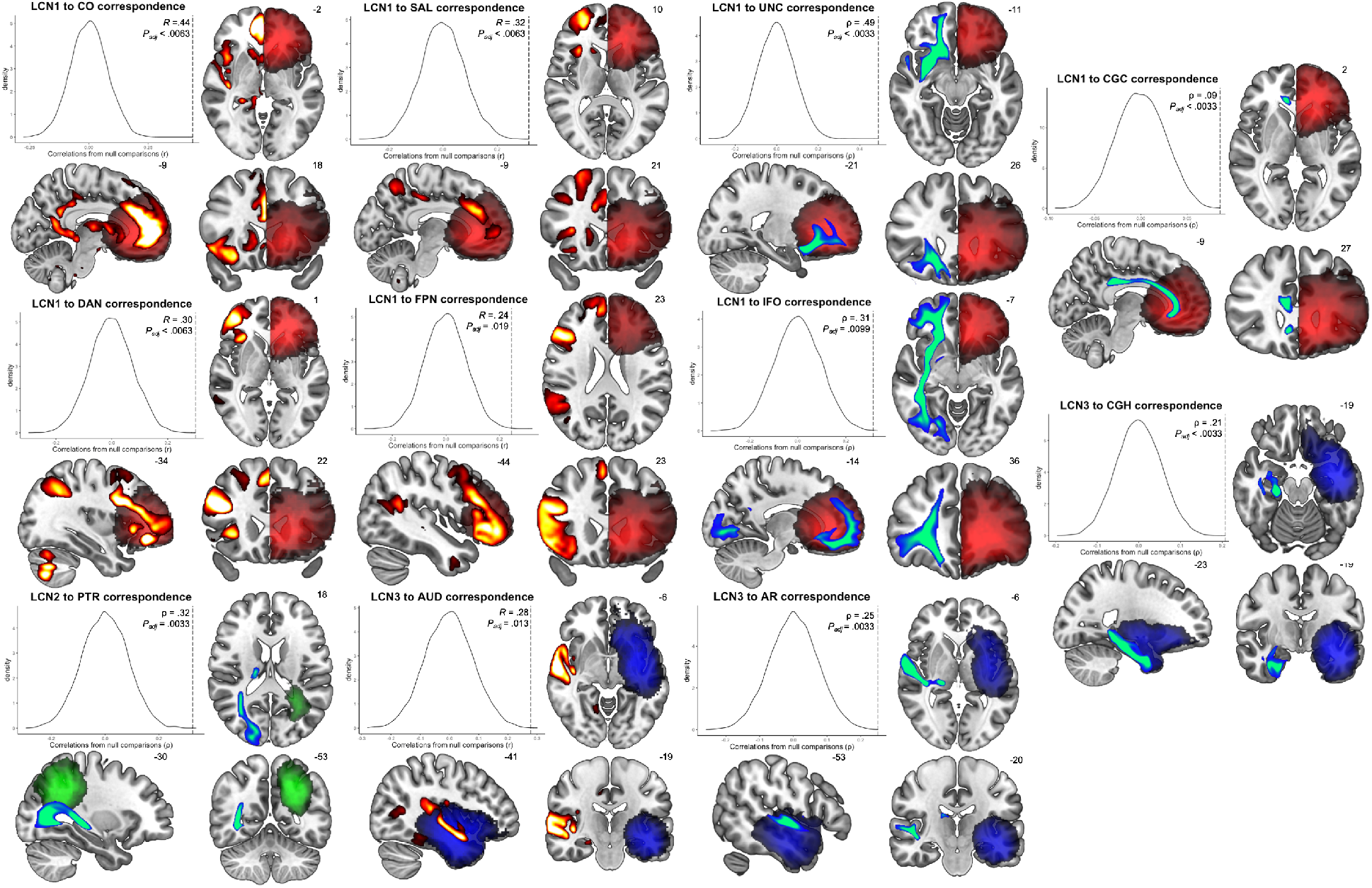
Functional and structural connectivity networks significantly associated with lesion covariance networks.

## Data Availability

Lesion data for GBM and LGG respectively are available at: https://wiki.cancerimagingarchive.net/display/DOI/Segmentation+Labels+and+Radiomic+Features+for+the+Pre-operative+Scans+of+the+TCGA-GBM+collection and https://wiki.cancerimagingarchive.net/display/DOI/Segmentation+Labels+and+Radiomic+Features+for+the+Pre-operative+Scans+of+the+TCGA-LGG+collection. Clinical and genomic data from The Cancer Genome Atlas can be downloaded in R by following the workflow described in https://www.bioconductor.org/packages/release/workflows/vignettes/TCGAWorkflow/inst/doc/TCGAWorkflow.html. UK BioBank neuroimaging data are available at: https://www.fmrib.ox.ac.uk/ukbiobank/.

https://wiki.cancerimagingarchive.net/display/DOI/Segmentation+Labels+and+Radiomic+Features+for+the+Pre-operative+Scans+of+the+TCGA-LGG+collection

https://www.bioconductor.org/packages/release/workflows/vignettes/TCGAWorkflow/inst/doc/TCGAWorkflow.html

https://www.fmrib.ox.ac.uk/ukbiobank/

## Ethics Statement

The research described in this manuscript used anonymized data made publicly available via The Cancer Genome Atlas and The Cancer Imaging Archive. All other data considered here had similarly been anonymized before being made publicly accessible.

